# Multi-omics integration identifies metabolic and inflammatory pathways underlying familial longevity

**DOI:** 10.64898/2026.05.12.26353027

**Authors:** Mengle Zhu, Niels van den Berg, Lieke Lamont, Esperanza Brashuis, Sabine Bos, Marian Beekman, Amy C. Harms, P. Eline Slagboom, Thomas Hankemeier, Joris Deelen

## Abstract

Familial longevity, quantified using the Longevity Relatives Count (LRC) score indicating the proportion of ancestral long-lived family members, associates with a pronounced 13 years delayed onset of cardiometabolic disease (CMD). Understanding the molecular basis of familial longevity therefore provides critical insights into mechanisms of cardiometabolic resilience. However, the combined metabolomics and proteomics profile associated with the delayed CMD onset observed in such long-lived family members is not understood yet. Hence, we integrated plasma metabolomics and proteomics in 495 participants from the Leiden Longevity Study to identify molecular signatures associated with (a contrast in) the LRC score. Metabolomics profiling captured 429 features, including amino acid derivatives, nucleosides, and lipid mediators, while proteomics quantified 374 proteins related to cardiovascular, metabolic, and inflammatory pathways. Three within-family analysis approaches were examined and overlapping findings were interpreted.

We identified ten metabolites and nine proteins that are associated with increased familial longevity, exemplified by a high LRC score. High LRC scoring individuals exhibited lower levels of amino acid derivatives (prolylhydroxyproline, 5-hydroxy-tryptophan, asymmetric dimethylarginine), nucleosides (2-methylguanosine, 7-methylguanosine, pseudouridine), N-acetylneuraminic acid and quinolinic acid, indicating optimized extracellular matrix integrity, vascular function, and reduced neuroinflammatory activity. Lipid mediators, including elevated 6-keto-PGF1a and reduced 9-HOTrE/alpha-linolenic acid ratio, reflected preserved endothelial homeostasis and attenuated inflammatory signaling. At the proteome level, strong ancestral familial longevity is associated with immune regulators (RETN, NPPB, IGSF8), extracellular matrix components (EFEMP1, EPHB4), and adhesion/signaling molecules (LRP11, ICAM3, KIT, ADGRG2), highlighting coordinated regulation of inflammation, tissue remodeling, and regenerative capacity.

Multi-omics pathway analyses indicated convergence on amino acid and nucleotide metabolism, lipid signaling, extracellular matrix remodeling, and receptor-mediated communication. Collectively, these multi-omics systemic signatures define a molecular framework of ancestral familial longevity characterized by reduced inflammation, preserved tissue integrity, and enhanced metabolic and regenerative processes. Our findings provide mechanistic insight into the biology of familial longevity and potentially cardiometabolic resilience.

## Introduction

The global demographic landscape is shifting dramatically, with the proportion of individuals aged 60 years or older projected to double by 2050 to nearly 2.1 billion. This makes population ageing a critical public health challenge worldwide [1]. However, the pace and quality of ageing strongly differ between individuals, reflecting complex interactions among genetic, molecular, social, behavioral, and environmental factors that influence biological ageing and disease susceptibility [2]. Understanding the mechanisms that provide protection from cardiometabolic and other chronic diseases to extend healthy lifespan (i.e. healthspan) is a primary goal of contemporary biogerontology [3, 4].

Studies of long-lived individuals have demonstrated that attaining a high age in the absence of chronic disease is possible and that this trait clusters in families [5–8]. We previously developed the Longevity Relatives Count (LRC) score to quantify familial longevity by calculating the proportion of an individual’s ancestors who belong to the top 10% longest-lived members of their birth cohort [9]. A high LRC score corresponds to low medication use and good metabolic and immune health in middle age, as indicated by MetaboHealth, a NMR metabolomics-based health biomarker [10]. Moreover, using data from the Leiden Longevity Study (LLS), we demonstrated that offspring of long-lived individuals exhibit a stunning cardiometabolic resilience illustrated by a 13 years delayed onset of the first cardiometabolic disease (CMD) in high LRC scoring offspring when compared to low LRC scoring controls [10]. This familial clustering of extended lifespan and healthspan suggests lower disease predisposition and/or the presence of inherited molecular mechanisms that confer protection against age-related traits [10]. Population-based studies of biological ageing and long-lived singletons (i.e. individuals without a known family history for longevity) suggest that distinct proteomic, metabolomic, and lipidomic profiles reflecting anti-inflammatory states, enhanced vascular function, and robust metabolic homeostasis mark healthy ageing [11–14]. However, the molecular mechanisms underlying delayed CMD onset in these long-lived family members have yet to be elucidated.

The ability to explore molecular networks underlying biological ageing and inherited longevity has been transformed by advances in high-throughput analytical technologies. Proteomics, particularly platforms such as Olink, enable quantitative profiling of hundreds of proteins implicated in inflammation, metabolism, and vascular signaling at high sensitivity and reproducibility [15, 16]. Metabolomics, based on techniques such as liquid chromatography-mass spectrometry (LC-MS), captures a broad array of low-molecular-weight metabolites and lipid mediators reflecting dynamic biochemical states [17, 18]. By integrating these complementary layers, i.e. proteomic, metabolomic, and clinical, multi-omics frameworks offer an unprecedented capacity to identify molecular axes associated with familial longevity and to characterize biological processes underlying ageing [19, 20].

In this study, we thus integrate proteomics and metabolomics data to characterize molecular signatures of LRC score-based familial longevity in participants of the LLS. Plasma samples were subjected to metabolomic and proteomic profiling, preprocessed for quality control and batch effects, and analyzed for associations with LRC-based familial longevity using three complementary analysis approaches. Significant molecular features were subsequently examined through (joint) pathway analyses to identify biological processes underlying familial longevity. This integrative framework provides new insights into molecular patterns linked to familial longevity and extends our understanding of the biological processes associated with healthy ageing (**Figure 1**).

**Figure 1.**
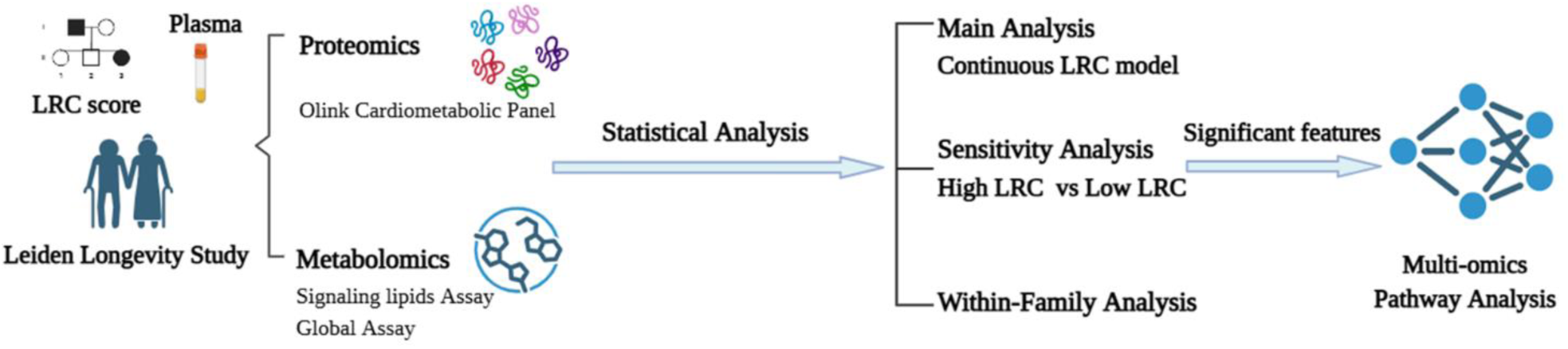
Study design overview. Plasma samples from Leiden Longevity Study participants were profiled by proteomics and metabolomics. The Longevity Relative Count (LRC) score was derived from ancestral lifespan data, and molecular features associated with LRC were identified through multiple analysis approaches and subjected to multi-omics pathway analysis to determine biological processes underlying familial longevity.

## Materials and Methods

### Study population

We used data from the Leiden Longevity Study (LLS); a family-based cohort initiated in 2002 to investigate mechanisms of healthy ageing and longevity [21]. In the LLS, 421 families were recruited with at least two long-lived siblings (F1 generation), defined as men aged ≥89 years and women aged ≥91 years. The study also enrolled the parents (F0 generation) and the offspring (F2 generation) of the long-lived sibling pairs, together with their partners, who served as household environment-matched controls. For the present analyses, we used the data of the third measurement timepoint of the participants (N=500), which took place between 2009 and 2010. Proteomic profiling was performed for all participants at this visit, whereas metabolomic profiling was available for a subset of participants (N = 333). The third visit of Leiden Longevity Study protocol was approved by the Medical Ethical Committee of the Leiden University Medical Center before the start of the study (P09.140) on the 5th of August 2009. The first participant was enrolled on the 24th of August 2009. In accordance with the Declaration of Helsinki, the Leiden Longevity Study obtained informed consent from all participants prior to their entering the study. Five participants were excluded due to missing LRC (longevity family recruitment criteria) information, resulting in an analytical dataset of 495 individuals for proteomic analyses. Of these, 329 individuals also had available metabolomic data and were included in the metabolomic analyses. Mortality information was verified using official records, including birth, marriage, and death certificates, as well as passports when available. In addition, verification took place via personal cards, which were obtained from the Dutch Central Bureau of Genealogy. In January 2025, all mortality information was updated through the Personal Records Database (PRD), which is managed by the Dutch governmental service for identity information (https://www.government.nl/topics/personal-data/personal-records-database-brp). The combination of officially documented information provides very reliable and complete ancestral as well as current mortality information.

### Longevity Relatives Count score

The Longevity Relatives Count (LRC) score, developed by van den Berg *et al.* [10], is a quantitative measure of familial longevity. The LRC score was used in LLS to map the offspring family history of longevity. The LRC score indicates the proportion of ancestors that became long-lived, weighted by the genetic distance between offspring and partners, and their ancestors. For example, an LRC of 0.5 indicates 50% long-lived ancestors. For this study, two generations of ancestors were available to calculate the LRC score for offspring and one generation for the partners of the LLS offspring. This captures the number of long-lived (survival to the 10% oldest survivors of a person’s own birth cohort and sex) family members of an individual.

### Metabolomics analysis

For metabolomics assessment, there was a selection of 333 EDTA plasma samples out of the total N=500 Leiden Longevity Study participants who visited LUMC between 2009 and 2010. Individuals were selected if they 1) were still alive in 2020, 2) had an APOE genotyping available, and 3) were >= 60 years of age.

Samples were stored at –80 °C since enrolment in 2009/2010 until analysis in 2024. Metabolite profiling was performed by Metabolomics and Analytics Centre of Leiden University using two mass spectrometry (MS)-based metabolomics assays: i) the global profiling assay [22] and ii) the signaling lipids assay [23]. The signaling lipids assay captures a wide range of lipid mediators, including various polyunsaturated fatty acids (PUFA) and their derived isoprostane and prostaglandin classes, specialized pro-resolving mediators (SPM), endocannabinoids, bile acids, sphingosine and sphinganine derivatives (including phosphorylated forms), and lysophospholipids. This assay is performed using liquid–liquid extraction followed by two separate chromatographic analyses: one under low pH and one under high pH conditions [23]. Quantification was performed using internal standard (ISTD) correction, and results were expressed as metabolite-to-ISTD ratios. The global profiling assay covers multiple compound classes, including acylcarnitines, amino acids and derivatives, indoles and derivatives, nucleosides and nucleotide analogues, phenols, and benzoic acid derivatives. This assay is performed using protein precipitation followed by LC-MS analysis in both positive and negative ionization modes [22], and results were reported as raw peak areas. A batch correction was applied to both assays based on quality control (QC) samples derived from pooled study plasma to minimize analytical drift. After quality control assessment using mzQuality [24], one sample was excluded due to an analytical error, resulting in 332 samples available for analysis. In addition, 169 signaling lipids and 153 global metabolites passed quality control and were retained for analyses.

From the measured metabolomics data, biologically relevant ratios were calculated to reflect enzyme-dependent conversions. Examples include ratios of secondary to primary bile acids, oxylipins to fatty acids, and various ratios among amines. This resulted in a comprehensive dataset of 429 metabolite features (169 signaling lipids and 153 global metabolites) and 107 calculated ratios (for the complete list, see **Table S1**).

For both assays, missing values, primarily due to concentrations below the limit of quantification, were first evaluated. Metabolites missing in more than 20% of samples within each assay were removed. Subsequently, the remaining metabolite values were log-transformed, and missing values were imputed using quantile regression imputation of left-censored data (QRILC). After imputation, both data sets were merged into a single data set. Finally, all metabolite features and ratios were subsequently Z-scaled to normalize distributions prior to downstream statistical analyses.

### Proteomics analysis

Proteomic analysis was performed on 500 EDTA plasma samples from the Leiden Longevity Study by the Genomics Core Facility (www.genomicserasmusmc.nl) in Rotterdam, following the manufacturer’s protocol [25]. Plasma samples were stored at –80 °C since enrolment in 2009/2010 until analysis in 2023. Quality control included exclusion of measurements below the limit of detection and assessment of inter-plate variability. Data were subsequently normalized and batch-corrected following Olink’s standard protocols. After quality control, all 374 proteins were retained for downstream analyses.

The Olink Explore assay (Olink, Uppsala, Sweden), based on the proximity extension assay technique, was used to quantify plasma levels of 374 proteins from the Cardiometabolic panel in the LLS. Data was transformed and normalized to Olink’s Normalized Protein eXpression (NPX) value. NPX is a relative protein quantification unit on a log2 scale, where a difference of 1 NPX equates to a doubling of protein concentration. NPX values are calculated from matched counts using Next-Generation Sequencing (NGS) as the readout, providing a standardized measure of protein abundance. The NPX values were then Z-scaled prior to statistical analysis.

### Statistical analysis

All statistical analyses were performed using R Studio version 4.4.2. All models were adjusted for age at inclusion and sex, which were selected a priori based on their known associations with metabolomic and proteomic profiles. Age and sex were self-reported, and body mass index (BMI; kg/m²) was calculated from measured height and weight and evaluated as a potential covariate. Medication use was obtained from pharmacy records covering the period 2006–2008 and categorized according to the Anatomical Therapeutic Chemical (ATC) classification system. Analyses focused on ATC groups A and C, specifically medications for diabetes (A10), hypertension (C02–C09), and hypercholesterolemia (C10). For each category, medication use was coded as a binary variable indicating whether participants had used at least one drug during the observation period.

To assess the relevance of body mass index (BMI) and medication use as covariates, their associations with the LRC score were evaluated prior to the main analyses. BMI or medication use were modeled as outcomes. Associations between medication use and the LRC score were examined using logistic regression models adjusted for age and sex, while the association between BMI and the LRC score was assessed using linear regression. Based on these analyses, medication use was included as a covariate in subsequent models, whereas BMI was not.

Associations between metabolite and protein levels and the LRC score were evaluated using regression models that account for family structure. Main and sensitivity analyses were performed using linear mixed-effects models with age, sex, and medication use as fixed-effect covariates, and family ID included as a random intercept to account for correlation of measurements within families. A mixed-effects model was chosen to properly account for the non-independence of related individuals and avoid biased estimates. Within-family analyses were conducted using a fixed-effects regression model comparing each offspring directly to their own partner, with partner ID capturing all shared household- or family-level effects. This approach provides a robustness check for genetically driven associations independent of shared environmental influences. Three complementary analyses were conducted: i) Main analysis (continuous LRC model): *Metabolite / Protein ∼ LRC_continuous_ + Age + Sex + Medication use + (1|Family ID)*. The LRC score was treated as a continuous variable to assess linear associations. ii) Sensitivity analysis (High LRC vs Low LRC model): *Metabolite / Protein ∼ LRC_binary_ + Age + Sex + Medication use + (1|Family ID)*. Offspring with a high LRC score (>0.3) were compared to partners with LRC = 0. This analysis was also used to explore potential non-linear effects, and iii) Within-family analysis: *Metabolite / Protein ∼ LRC_continuous_ + Age + Sex + Medication use, with family fixed effects(Partner ID)*. In this analysis, 247 offspring were directly compared to their own partner to minimize environmental variation.

Effect sizes and statistical significance were reported for all metabolites and proteins across the continuous LRC, high LRC vs low LRC, and within-family analyses. Multiple testing correction (FDR) was applied using the Benjamini-Hochberg (BH) method to control for false positives.

### Multi-omics pathway enrichment analysis

Differentially expressed proteins (DEPs) and differential metabolites (DMs) across different analysis methods were identified using the same selection criteria (p-values < 0.05). For Metabolomics data, pathway enrichment analysis of DMs was performed using MetaboAnalyst (version 6.0) according to the KEGG database. Pathways with a p-value <0.05 were considered nominally enriched and interpreted as exploratory findings. For Proteomics data, enrichment analysis for DEPs was conducted using STRING (version 12.0) according to the Gene Ontology (GO) database. Pathways with p_FDR_ < 0.05 were considered significant. To better understand the interactions between proteins and metabolites, identified DEPs and DMs were subjected to joint pathway analysis using MetaboAnalyst based on the KEGG database. The impact score is calculated based on the position of significant metabolites or genes (hits) within each pathway. A high-impact pathway indicates that hits occur at key positions, suggesting that even small changes may strongly influence pathway function. Conversely, a low-impact pathway indicates that hits are located on peripheral nodes, so biological significance may be limited even if the enrichment p-value is low. Pathways with an impact > 0.10 and p-value < 0.05 were considered significant.

## Results

### Study populations

Baseline characteristics of the study participants included in proteomics (n=495) and metabolomics (n=329) analyses are summarized in **Table 1**. Participants had a mean age of 65.9 years (range 45.5-84.8) in the proteomics dataset and 67.7 years (range 61.0-84.8) in the metabolomics dataset. Approximately half of the participants were female in both datasets (50.5% and 50.6%, respectively). Mean BMI was similar across datasets (25.6 kg/m² in both proteomics and metabolomics), just above the threshold for overweight, with values ranging from 17.6 to 44.6 kg/m². Information on medication use was obtained from participants’ pharmacy records. In the proteomics and metabolomics datasets, 21.6% and 24.3% of participants, respectively, were taking antihypertensive medications; 12.3% and 15.5% were on lipid-lowering therapy; and 4.2% and 5.2% were receiving antidiabetic treatment. Regarding longevity family status, 273 and 194 participants had a high LRC score (>0.3) and 183 and 109 in the proteomics and metabolomics datasets respectively, while 183 and 109 participants, respectively, had an LRC score of 0. After accounting for family structure, 247 couples were available for within-family proteomics analyses and 128 couples for within-family metabolomics analyses. A total of 374 proteins were quantified in the proteomics analysis, and 429 metabolic features were measured in the metabolomics analysis.

**Table 1.** Baseline characteristics of participants included in metabolomics and proteomics analyses from the Leiden Longevity Study.

| Variable | Proteomics (N=495) | Metabolomics (N=329) |
| --- | --- | --- |
| **Demographics** |  |  |
| Females, n (%) | 250 (50.5%) | 166 (50.6%) |
| Age, mean (range), years | 65.9 (45.5-84.8) | 67.7 (61.0-84.8) |
| BMI, mean(range), kg/m<sup>2</sup> | 25.6 (17.6-44.6) | 25.6 (18.3-44.6) |
| **Medication use, n (%)** |  |  |
| Antidiabetic drugs | 21 (4.2%) | 17 (5.2%) |
| Antihypertensive drugs | 107 (21.6%) | 80 (24.3%) |
| Lipid-lowering drugs | 61 (12.3%) | 51 (15.5%) |
| **Family status** |  |  |
| High LRC (>0.3) | 273 | 194 |
| Low LRC (0) | 183 | 109 |
| Within-family couples, n (female %) | 247 (50.0%) | 128 (50.0%) |
| **Omics features, n** | 374 | 429 |

Values are mean (range) or number (%). BMI: body mass index; LRC: Longevity Relatives Count score. Within-family couples represent offspring-partner pairs included for matched analyses.

### Associations of medication use and BMI with LRC score

Associations of BMI and medication use with the LRC score were evaluated to determine their relevance as covariates in subsequent analyses. Logistic regression analyses indicated that a higher LRC score is associated with lower odds of medication use. Specifically, a 10% increase in the LRC score was associated with a 10% reduced odds of using antihypertensive drugs (OR = 0.90, 95% confidence interval (CI): 0.90–0.91), 19% reduced odds of using lipid-lowering drugs (OR = 0.81, 95% CI: 0.71–0.94), and 19% reduced odds of using antidiabetic drugs (OR = 0.81, 95% CI: 0.66–0.99), after adjusting for age and sex. Linear regression analysis showed that BMI was not significantly associated with the LRC score (β = –0.07, 95% CI: –0.20 to 0.05). Full results are provided in **Table S2**. Based on these findings, medication use was included as a covariate in subsequent analyses, whereas BMI was not.

### Associations of metabolites with familial longevity

#### Associations of metabolites with continuous LRC score

To identify metabolites associated with the LRC score, a linear mixed-effects regression analysis was performed, adjusting for age, sex, medication use, and family ID. Twenty-six differential metabolites (DMs) were nominally significantly associated with the LRC score (p < 0.05; **Table S3**). However, none of these associations was significant after FDR adjustment for multiple testing. Higher levels of amino acid-related metabolites (e.g. prolylhydroxyproline and 5-hydroxy-tryptophan), nucleosides (e.g. 7-methylguanosine and 2-methylguanosine), and markers of inflammation and oxidative stress (e.g. N-acetylneuraminic acid and the ratio of 9-HOTrE to alpha-linolenic acid (aLA)) were associated with lower LRC score. Conversely, higher levels of lipid mediators (e.g. 11b-PGE2 and 6-keto-PGF1a) were associated with higher LRC score.

#### Associations of metabolites with high and low LRC scoring groups

To complement the primary analysis of all 329 participants, a sensitivity analysis was performed, including only individuals with a high LRC score (>0.3; n = 194) and those with an LRC score of 0 (n = 109). Using the same linear mixed-effects regression framework but treating LRC score as a binary variable, 23 DMs were nominally significantly associated with the LRC group status (**Table S4**). However, none of these associations was significant after FDR adjustment for multiple testing. Among these, 19 overlapped with findings from the continuous LRC score analysis, showing consistent directions of effect. These included amino acid-related metabolites, nucleotide derivatives, lipid mediators, as well as malic acid and N-acetylneuraminic acid. Most metabolites were higher in participants with the lower LRC score, except 6-keto-PGF1α. In addition, four metabolites, dopamine, N2,N2-dimethylguanosine, uric acid, and 7-HDoHE, were uniquely identified in this sensitivity analysis, all of which were present at lower levels in the high LRC group.

#### Associations of metabolites with the LRC score within the family

The original design of the LLS was meant to minimise household-related environmental influences on familial longevity by comparing offspring of long-lived individuals with their own partners. Therefore, a within-family analysis was conducted to complement the LRC score analyses, aiming to distinguish associations driven by familial longevity from those related to shared lifestyle factors. After excluding 72 participants without matching pairs, 128 offspring-partner pairs remained. Analyses were adjusted for age, sex, and medication use.

In total, 38 DMs were nominally significantly associated with the LRC score (**Table S5**). However, none of these associations remained significant after FDR correction for multiple testing. Of these, 26 metabolites were negatively associated with the LRC score, showing lower levels in participants with higher LRC score. These included several nucleosides (e.g. N2,N2-dimethylguanosine and pseudouridine), amino acid derivatives (e.g. N-acetylserine, prolylhydroxyproline and asymmetric dimethylarginine), and other metabolites (e.g. uric acid and N-acetylneuraminic acid), as well as lipid- and bile acid-related metabolites (e.g. chenodeoxycholic acid (CDCA) and the 9-HOTrE/aLA ratio). Conversely, 12 metabolites were positively associated with the LRC score, showing higher levels in participants with higher LRC score, including lysophospholipids (e.g. LPA16:0, LPA18:1, LPE18:0), lipid mediators (e.g. 5,6-DiHETrE, 6-keto-PGF1a), and several bile acid ratios (e.g. TLCA/LCA and TCDCA/CDCA).

#### Summary of nominally significant metabolites across analyses

To integrate findings from all three analytical models, effect size estimates (β) and 95% confidence intervals for 56 nominally significant metabolites were examined (**Figure 2**). Ten metabolites were identified as nominally significant across the continuous, high vs. low, and within-family analyses, indicating robust associations with familial longevity. Across the three analytical approaches, we observe substantial overlap and consistent effect directions. Three amino acid derivatives (prolylhydroxyproline, 5-hydroxy-tryptophan, and asymmetric dimethylarginine), three nucleosides (7-methylguanosine, 2-methylguanosine, and pseudouridine), one additional metabolite (quinolinic acid), and two markers of inflammation or oxidative stress (N-acetylneuraminic acid and ratio of 9-HOTrE to aLA) generally showed lower levels with higher LRC score, whereas lipid mediators (e.g. 6-keto-PGF1a) exhibit the opposite pattern. Collectively, these results indicate a robust metabolic signature of familial longevity emerging from three analysis approaches, characterized by coordinated alterations in amino acid derivatives, nucleosides, inflammatory markers, and lipid mediators.

**Figure 2:**
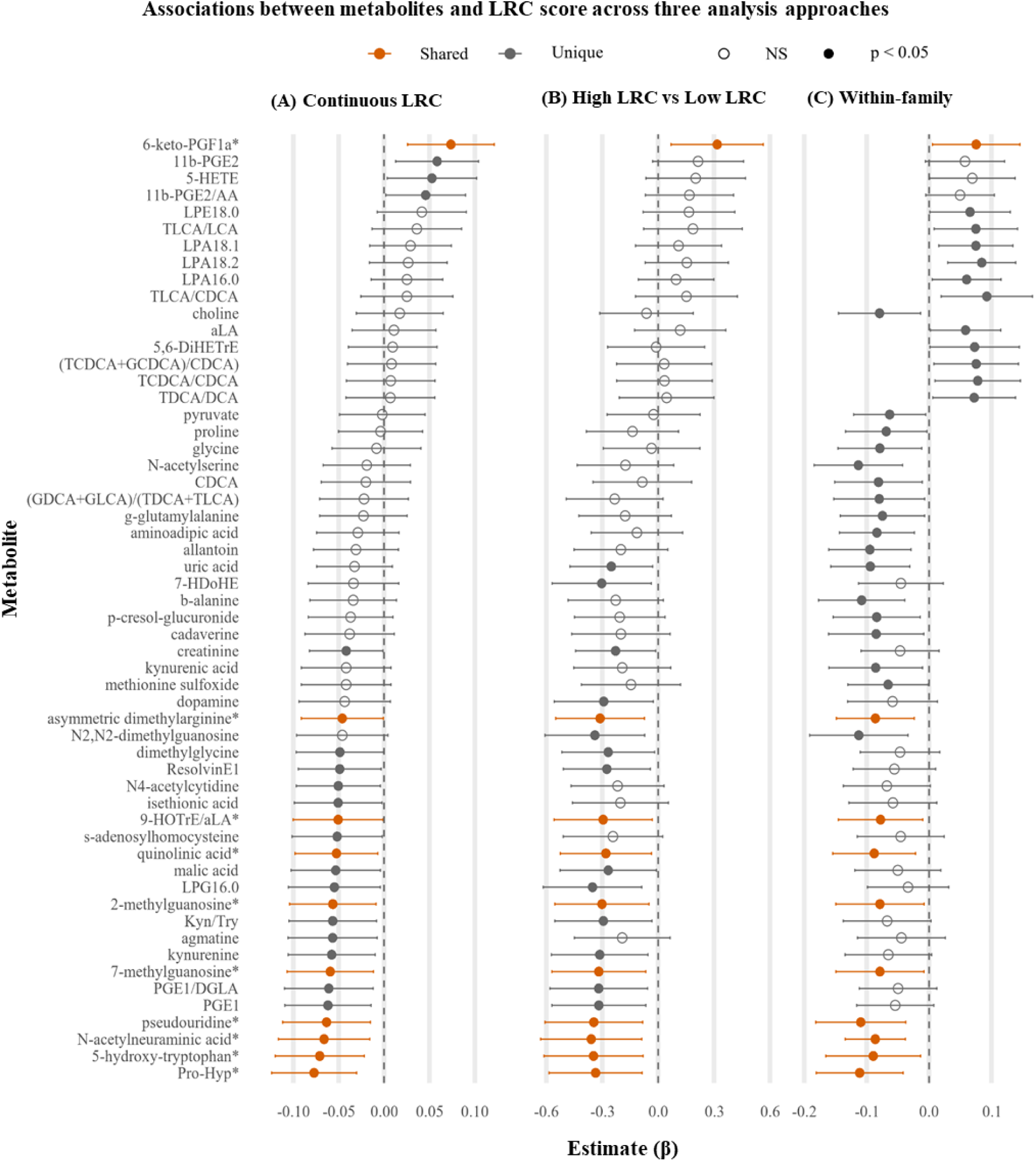
Associations between metabolites and LRC score across three analysis approaches: (A) Continuous LRC, (B) High LRC vs Low LRC and (C) Within-family. Points represent effect size estimates (β) with 95% confidence intervals; filled points indicate p < 0.05, open points indicate non-significant (NS) results. Orange points indicate metabolites are significant across all three approaches (marked with an asterisk * in the plot), while grey points indicate metabolites unique to a single analysis. The dashed vertical line indicates the null value (β = 0). Abbreviations: Pro-Hyp: prolylhydroxyproline; aLA: alpha-linolenic acid; Kyn/Try: ratio of kynurenine/tryptophan.

### Associations of proteins with familial longevity

#### Associations of proteins with continuous LRC score

Linear mixed-effects regression adjusted for age, sex, medication use, and family ID identified 22 differentially expressed proteins (DEPs) nominally significantly associated with the LRC score (p < 0.05; **Table S6**). However, none of these associations was statistically significant after FDR adjustment. Seventeen DEPs show lower levels in the individuals with higher LRC score, including immune-related proteins (e.g. CHIT1 and IGSF8), adipokines/cardiovascular markers (e.g. RETN, NPPB, TCL1B), extracellular matrix proteins (e.g. EFEMP1 and EPHB4), and adhesion and signaling molecules (e.g. ICAM3 and LRP11). Conversely, five DEPs have higher levels in the participants with higher LRC score. This includes adhesion and signaling molecules (e.g. KIT, ADGRG2 and GRK5), and metabolic/oxidative stress-related enzymes (e.g. GLRX and AKR1C4).

#### Associations of proteins with high and low LRC scoring groups

In addition to the continuous LRC score analysis, individuals with a high LRC score (>0.3, n = 273) were compared with those with an LRC score of 0 (n = 183). This analysis identified 20 DEPs that were nominally significantly associated with LRC group status (**Table S7**). However, these associations were not significant after FDR adjustment. Of these, 15 overlap with the proteins identified in the continuous model and exhibited consistent directions of effect, reinforcing the robustness of associations.

Five additional DEPs are uniquely identified in this comparison. Four DEPs, NT-proBNP (a cardiac biomarker), LEP (leptin, an adipokine), SOST (sclerostin, bone metabolism), and CPA1 (carboxypeptidase A1, pancreatic enzyme), show lower levels in the high LRC group. In contrast, DPP4 (dipeptidyl peptidase-4, an enzyme involved in glucose metabolism and immune regulation) show higher levels in the high LRC group.

#### Associations of proteins with LRC score within the family

To account for shared environmental factors, a within-family analysis was conducted among 247 offspring-partner pairs, adjusting for age, sex, and medication use. Twenty-six DEPs are nominally significantly associated with the LRC score (**Table S8**). However, none of the associations remained significant following FDR correction. Twenty proteins show lower levels in the participants with higher LRC score, including markers of cardiovascular stress (e.g. NPPB and MB), extracellular matrix and cytoskeletal proteins (e.g. CTSB and EFEMP1), and metabolic/inflammatory regulators (e.g. RETN and IGSF8). Six DEPs show higher levels in the individuals with higher LRC score, notably including growth factor and signaling proteins (e.g. NTRK2 and KIT) and renal/metabolic regulators (e.g. APLP1 and ADGRG2).

#### Summary of nominally significant proteins across analyses

To summarize findings across all three analytical approaches, a forest plot was generated (**Figure 3**), with Panels A-C representing the continuous LRC, high LRC vs low LRC, and within-family analyses. Effect size estimates (β) with 95% confidence intervals are shown for all 40 nominally significant proteins. Nine proteins, marked with an asterisk (*), are nominally significant across all analyses and are highlighted in orange to emphasize robust results.

**Figure 3.**
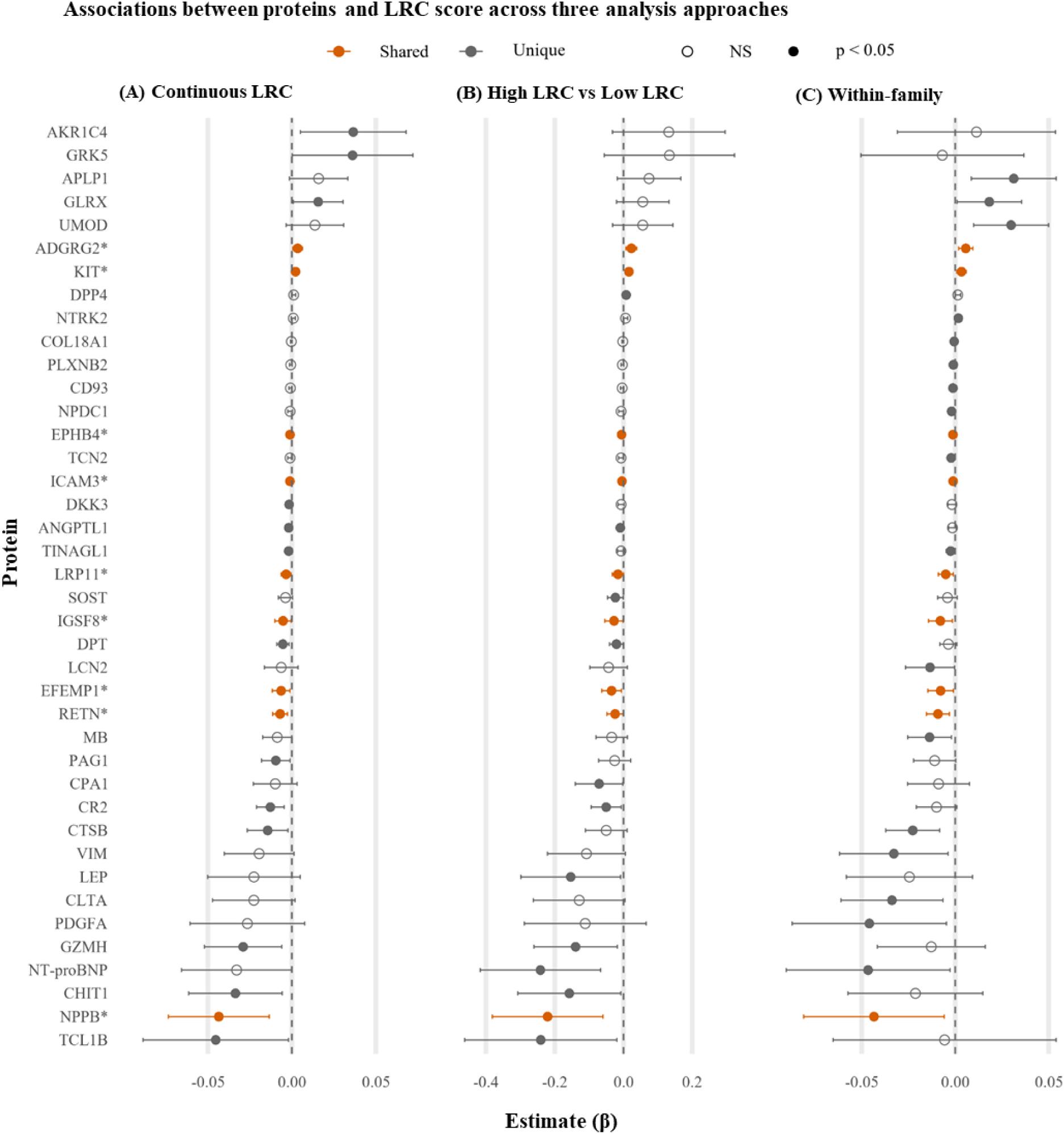
Associations between proteins and LRC score across three analysis approaches: (A) Continuous LRC, (B) High LRC vs Low LRC, and (C) Within-family. Points represent effect size estimates (β) with 95% confidence intervals; filled points indicate p < 0.05, open points indicate non-significant (NS) results. Orange points indicate metabolites are significant across all three approaches (marked with an asterisk * in the plot), while grey points indicate metabolites unique to a single analysis. The dashed vertical line indicates the null value (β = 0).

Shared proteins included immune and inflammatory regulators (RETN, NPPB, IGSF8), extracellular matrix components (EFEMP1, EPHB4), and membrane-associated adhesion and signaling molecules (LRP11, ICAM3, KIT, ADGRG2). The observed concordance across all analytical approaches supports a robust proteomic signature linked to familial longevity, characterized by coordinated regulation of immune responses, extracellular matrix components, and receptor-mediated signaling involved in cellular homeostasis.

### Multi-omics pathway analysis of proteins and metabolites associated with familial longevity

To gain deeper insight into the biological processes underlying familial longevity, pathway enrichment analysis was performed on the 40 significant DEPs and 56 significant DMs identified across three analysis approaches in the study (**Table S9**). Analyses were conducted using Gene Ontology (GO) and Kyoto Encyclopedia of Genes and Genomes (KEGG) in *MetaboAnalyst* and *STRING*, enabling identification of molecular networks and biological pathways associated with longevity.

Metabolomic pathway enrichment analysis (**Figure 4A**) identified nine nominally significantly enriched metabolic pathways (p < 0.05). These included glycine, serine and threonine metabolism, primary bile acid biosynthesis, glutathione metabolism, glyoxylate and dicarboxylate metabolism, arginine and proline metabolism, biosynthesis of unsaturated fatty acids, tryptophan metabolism, arachidonic acid metabolism, and the citrate cycle (TCA cycle) (**Table S10**).

**Figure 4.**
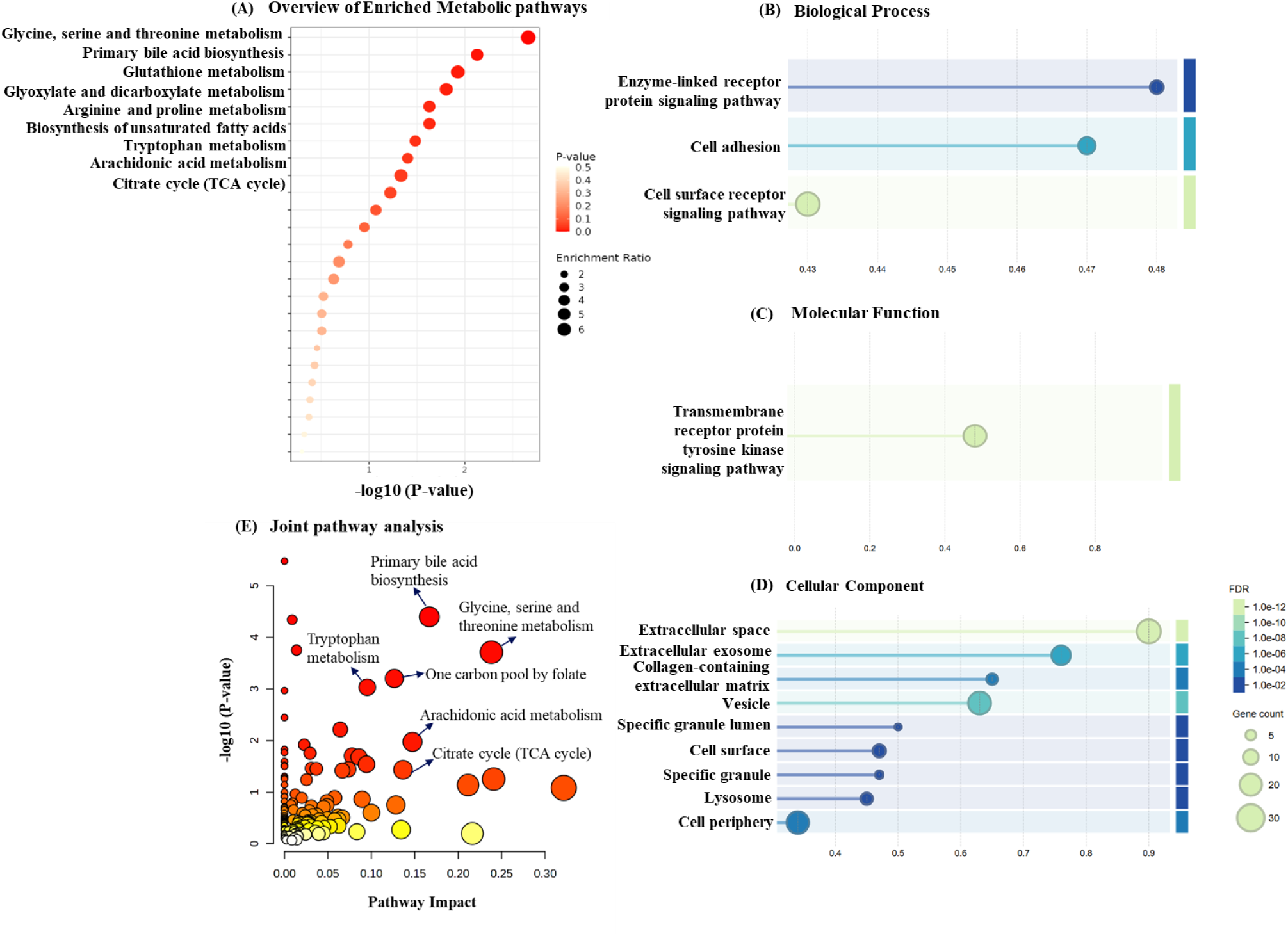
Pathway analysis of significant metabolites and proteins. (A). KEGG-based metabolic pathway enrichment of significant metabolites. Gene Ontology (GO) enrichment of significant proteins across three categories: (B) Biological Process, (C) Cellular Component, and (D) Molecular Function. (E). Integrated pathway analysis combining significant metabolites and proteins. The size and color of the bubbles represent the enrichment factor and the p-value, respectively. Darker colors indicate more significant p-values, and larger bubbles indicate higher impact scores.

Notably, several key metabolites central to our findings were not captured by automated analysis due to missing KEGG identifiers or incomplete pathway annotations. To address this, ten shared metabolic features were manually mapped to their relevant biological pathways. These included intermediates of the tryptophan-kynurenine axis (5-hydroxy-tryptophan, quinolinic acid), markers of lipid mediator turnover (9-HOTrE/aLA ratio, 6-keto-PGF1a), components of arginine/proline metabolism (prolylhydroxyproline, asymmetric dimethylarginine), which are linked to collagen turnover, nitric oxide synthesis, and vascular function, and metabolites related to nucleotide modification pathways (7-methylguanosine, 2-methylguanosine, pseudouridine, N-acetylneuraminic acid). Collectively, these results indicate coordinated regulation of amino acid, lipid, and nucleotide metabolism in long-lived families.

GO enrichment analysis of DEPs identified several significantly affected biological processes (p_FDR_ < 0.05), including the enzyme-linked receptor protein signaling pathway, cell adhesion, and cell surface receptor signaling pathways (**Figure 4B**). Molecular function enrichment was observed in transmembrane receptor protein tyrosine kinase signaling (**Figure 4C**). At the cellular component level, enrichment was observed in multiple compartments, including the extracellular region (extracellular space, exosomes, collagen-containing extracellular matrix), vesicular structures (vesicles, lysosomes, specific granule lumen, specific granule), and the plasma membrane region (cell periphery and cell surface) (**Figure 4D**). Shared proteins such as KIT, ADGRG2, EPHB4, ICAM3, LRP11, IGSF8, EFEMP1, RENT, and NPPB were predominantly associated with receptor signaling, cell adhesion, and extracellular communication (**Table S11**).

Integrated proteomic-metabolomic analysis confirmed overlapping pathways between DEPs and DMs (**Figure 4E**). Primary bile acid biosynthesis showed the most significant enrichment, while glycine, serine, and threonine metabolism had the highest pathway impact. Additional pathways enriched in the integrated analysis included tryptophan metabolism, one-carbon pool by folate, arachidonic acid metabolism, and the TCA cycle (**Table S12**). Although not directly linked mechanistically, the involvement of arginine-proline metabolism (e.g., prolylhydroxyproline, asymmetric dimethylarginine) and EFEMP1 suggests coordinated extracellular matrix remodeling, integrating collagen turnover with matrix structural organization.

Collectively, these results demonstrate that proteomic and metabolomic alterations associated with familial longevity converge on interconnected metabolic and signaling networks, particularly amino acid metabolism (tryptophan, arginine/proline), lipid mediator turnover (arachidonic acid), and extracellular receptor signaling, suggesting coordinated regulation of metabolic homeostasis and intercellular communication.

## Discussion

In this study, metabolomic and proteomic systemic signatures associated with LRC score-based ancestral familial longevity were investigated in the Leiden Longevity Study (LLS) cohort. By exploring features that were consistent across three analysis approaches, including a continuous, contrasting high vs low, and within-family analyses of the LRC score, we identified robust associations for ten metabolites and nine proteins. These molecular signatures converge on amino acid and nucleotide metabolism, lipid signaling, extracellular matrix remodeling, and receptor-mediated signaling. This suggests that the systemic basis of familial longevity and the cardiometabolic resilience that long-lived families from the LLS express entails a coordinated regulation of metabolic, inflammatory, intercellular and tissue remodeling pathways.

The metabolomic analyses highlighted amino acid derivatives, nucleosides, and lipid mediators as key systemic signatures of familial longevity (**Figure 2**). Specifically, amino acid-related metabolites such as prolylhydroxyproline, 5-hydroxy-tryptophan, and asymmetric dimethylarginine were lower in individuals with higher LRC score, suggesting optimized regulation of extracellular matrix remodeling, vascular homeostasis, and nitric oxide signaling, pathways central to cardiovascular and tissue health. Reduced prolylhydroxyproline may reflect diminished collagen turnover and greater matrix stability, which could help maintain vessel elasticity, reduce fibrosis, and support tissue repair over time [26, 27]. Lower asymmetric dimethylarginine levels indicate enhanced endothelial function and improved nitric oxide bioavailability, promoting healthy vascular tone and perfusion [28, 29]. Lower levels of 5-hydroxy-tryptophan may contribute to balanced serotonin metabolism, which could influence vascular tone, neurovascular communication, and metabolic homeostasis [30]. Consistent with these patterns, lower levels of quinolinic acid, a downstream metabolite of the tryptophan-kynurenine pathway, suggest reduced neuroinflammatory signaling, potentially limiting microglial activation, reducing excitotoxicity, and supporting neuronal health and cognitive function with ageing [31]. Additional reductions in modified nucleosides, including 7-methylguanosine, 2-methylguanosine, and pseudouridine, may reflect more efficient RNA turnover and diminished cellular stress, thereby enhancing proteostasis, maintaining protein quality control, and supporting systemic metabolic efficiency [32]. Lower levels of N-acetylneuraminic acid (sialic acid) similarly align with reduced systemic inflammation and improved endothelial and cognitive function, complementing findings that elevated sialic acid is linked to cardiometabolic dysfunction [33, 34].

Distinct lipid signatures further characterized the longevity-associated metabolome. A reduced 9-HOTrE/aLA ratio suggests lower lipid peroxidation and attenuated pro-inflammatory signaling, potentially protecting the endothelium and reducing oxidative stress-related tissue damage [35]. In contrast, elevated 6-keto-PGF1a, a stable prostacyclin metabolite, reflects preserved endothelial function, antithrombotic activity, and regulation of vascular tone, supporting cardiovascular health and long-term tissue perfusion [36, 37]. Together, these lipid mediators integrate with amino acid- and nucleotide-related changes to suggest a metabolic phenotype that preserves structural integrity, enhances tissue perfusion, and increases resilience to age-related vascular damage. In summary, the metabolomic profile associated with familial longevity highlights coordinated regulation of amino acid, nucleotide, and lipid metabolism that collectively support vascular integrity, inflammation control, and cellular resilience. These molecular mechanisms likely underpin the extended healthspan observed in longevity-enriched families, linking genetic propensity for longevity to measurable physiological outcomes.

Proteomic analysis identified nine proteins that associate with familial longevity in all three analytical approaches: RETN, NPPB, IGSF8, EFEMP1, ICAM3, EPHB4, KIT, ADGRG2, and LRP11 (**Figure 3**). These proteins converge on pathways regulating inflammation, extracellular matrix remodeling, vascular homeostasis, and intercellular signaling, reflecting systemic adaptations that support resilience in ageing. Lower levels of RETN, NPPB, and IGSF8 in individuals with higher LRC score suggest reduced chronic inflammation and improved cardiometabolic health. Reduced NPPB (B-type natriuretic peptide) aligns with diminished cardiac stress and favorable cardiovascular outcomes observed in long-lived populations [38], while decreased RETN (resistin) suggests lower vascular inflammation and improved insulin sensitivity [39, 40]. Lower IGSF8 (Immunoglobulin superfamily member 8), an immune checkpoint regulator that modulates natural killer cell activity, may reflect enhanced immune surveillance and reduced systemic immune activation, hallmarks of healthy ageing [41]. Complementing these inflammatory markers, extracellular matrix-related proteins EFEMP1 (Fibulin-3) and EPHB4 were lower in high-LRC individuals, consistent with their established associations with vascular dysfunction, cellular senescence, and cancer progression [42–46], and suggesting that moderated extracellular remodeling contributes to structural tissue stability and long-term vascular integrity.

Adhesion and receptor signaling proteins, including LRP11, ICAM3, KIT and ADGRG2, further highlight a balance between immune regulation and regenerative capacity. LRP11, a membrane-associated receptor involved in lipid transport and oncogenic signaling, was lower in high-LRC individuals, indicating reduced metabolic and cancer-related stress [47, 48]. ICAM3, a leukocyte adhesion molecule, was also reduced, reflecting a tempered inflammatory state [49]. In contrast, KIT, a receptor tyrosine kinase essential for stem cell maintenance, tissue regeneration, and mitochondrial function [50, 51], and ADGRG2, an adhesion G protein-coupled receptor involved in tissue repair and immune modulation [52, 53], were elevated in high-LRC participants. This coordinated expression pattern, reduced ICAM3 and LRP11 together with elevated KIT and ADGRG2, suggests an adaptive shift favoring efficient regeneration of vascular, connective, and immune tissues while maintaining controlled immune signaling, processes critical for sustaining physiological integrity in long-lived families. Overall, the proteomic signatures of familial longevity point to integrated mechanisms of reduced inflammation, preserved extracellular and vascular integrity, and enhanced regenerative and immune-regulatory capacity, which likely support systemic homeostasis and contribute to the extended healthspan observed in long-lived families.

Our findings can be contextualized within previous investigations of familial longevity, although direct multi-omic comparisons remain limited. The Long Life Family Study (LLFS) assessed metabolomic profiles in offspring of long-lived families, identifying signatures linked to cardiovascular and metabolic health that partially align with our observations of amino acid, nucleotide, and lipid metabolism modulation [54]. Similarly, the New England Centenarian Study (NECS) measured metabolomics and proteomics in centenarians and their offspring, highlighting altered inflammatory, extracellular matrix, and tryptophan-kynurenine pathways, consistent with our findings and suggesting shared mechanisms in neurovascular and metabolic resilience [55–57]. The MARK-AGE study, a European cohort investigating ageing biomarkers, did not include proteomic or metabolomic analyses in offspring of long-lived individuals but underscores the value of integrative biomarker approaches for ageing research [58]. Extending beyond familial cohorts, large non-family-based studies have also reported metabolic and proteomic signatures linked to longevity. Plasma proteomic analyses highlighted pathways related to inflammation, cardiovascular homeostasis, and extracellular matrix regulation [59], while a large-scale metabolomic study revealed alterations in nucleotide and lipid metabolism, consistent with our observations of coordinated metabolic regulation [32]. Notably, recent proteomic analyses within the Leiden Longevity Study identified circulating proteins associated with mid- and late-life survival, cardiometabolic healthspan, and enrichment in longevity families [60]. In the present study, we deliberately applied the Olink platform, capturing a distinct set of circulating proteins, and integrated these data with metabolomic profiling to investigate signatures of familial longevity. Although there was no overlap at the level of individual protein markers, the combined proteomic and metabolomic signatures converge on biological processes related to inflammation and metabolic regulation, in line with this previous study. Collectively, these studies, together with our multi-omic analysis, highlight convergent and complementary molecular mechanisms underpinning familial longevity, while emphasizing the need for standardized, cross-cohort methodologies to enable replication and direct comparisons.

While these previous studies provide valuable context and highlight shared biological pathways, our integrated multi-omic analysis offers unique insights into the coordinated regulation of metabolism, extracellular matrix remodeling, and receptor-mediated signaling in familial longevity. Despite these advances, direct molecular overlaps between the proteomic and metabolomic layers in our cohort were modest. This modest intersection likely reflects several factors. Metabolites capture rapid biochemical fluxes, whereas proteins primarily reflect more stable structural and signaling states, with enzymes mediating the underlying reactions. The targeted proteomic panel focused primarily on cardiovascular-related proteins, which may have missed other health-related pathways. Incomplete metabolite annotation, including missing KEGG identifiers for several key features, further constrained direct cross-omic mapping. In addition, the moderate sample size may have limited power to detect subtle effects, and no single metabolite or protein association remained statistically significant after FDR correction. Finally, these findings are based on a single study, as no other cohorts currently provide the multigenerational data required to calculate the LRC score, limiting opportunities for independent replication. Future studies leveraging larger, multi-ethnic cohorts and comprehensive multi-omics integration, including transcriptomics, epigenomics, and single-cell resolution approaches, will be essential to further delineate molecular networks underlying familial longevity and extended healthspan.

In conclusion, our study successfully identified robust molecular signatures associated with familial longevity. Coordinated regulation of amino acid and nucleotide metabolism, lipid signaling, extracellular matrix remodeling, and receptor-mediated communication collectively promotes reduced inflammation, preserved vascular and tissue integrity, and optimized metabolic function. A key strength of this work is the use of three complementary analytical approaches, namely continuous, high vs low, and within-family comparisons, to robustly assess familial longevity. Furthermore, the availability of matched metabolomic and proteomic data in the same participants allowed an integrated, multi-omic characterization of molecular pathways. Importantly, these findings provide a more detailed and mechanistic view of inflammatory regulation and tissue regeneration than traditional cytokine-based measures, uncovering molecular pathways of familial longevity that were previously uncharacterized. By linking genetic propensity for longevity, as quantified by the LRC score, to specific molecular profiles, this work advances our understanding of how heritable factors translate into sustained physiological health and extended healthspan.

## Author contributions

M.Z.: Conceptualization, investigation, methodology, data curation, formal analysis, visualization, writing − original draft. N.V.D.B.: Conceptualization, investigation, methodology, visualization, writing − original draft. L.L.: Conceptualization, investigation, methodology, visualization, supervision, writing − review & editing. E.B.: Investigation, writing − review & editing. S.B.: Investigation, writing − review & editing. M.B.: Conceptualization, visualization, resources, writing − review & editing. A.H.: Supervision, writing − review & editing. P.E.S.: Resources, funding acquisition, writing − review & editing. T.H.: Supervision, funding acquisition, writing − review & editing. J.D.: Conceptualization, investigation, methodology, visualization, supervision, writing − review & editing.

## Data Availability

The LLS data are protected by Dutch personal integrity laws and other (privacy) regulations and are thus not publicly available. However, most of the individual-level data are accessible following our data access procedure (https://leidenlangleven.nl/data-access/). Summary statistics belonging to the current manuscript are available upon request to the corresponding author Joris Deelen.

## Acknowledgements

This project received funding from the China Scholarship Council (no. 202107060001) and the Vitality Oriented Innovations for the Lifecourse of the Ageing Society (VOILA) project funded by ZonMw (no. 457001001). Additional support was provided by the Netherlands X-omics Initiative (NWO Project No. 184.034.019) and the project Building the Infrastructure for Exposome Research: Exposome-Scan (Project No. 175.2019.032), funded under the “Investment Grant NWO Large” program of the Dutch Research Council (NWO). Additionally, we would like to thank Alida Kindt (Leiden University) for her valuable guidance and suggestions regarding data analysis in this study.

